# Vaccine hesitancy, reactogenicity and immunogenicity of BNT162b2 and CoronaVac in pediatric patients with neuromuscular diseases

**DOI:** 10.1101/2022.11.29.22282857

**Authors:** Michael Kwan Leung Yu, Hoi Shan Sophelia Chan, Samuel Cheng, Daniel Leung, Sau Man Chan, Amy Ka Yan Suen, Wilfred Hing Sang Wong, Malik Peiris, Yu Lung Lau, Jaime S Rosa Duque

## Abstract

**Introduction:** COVID-19 causes global health and psychosocial devastation, particularly to high-risk patients such as those with neuromuscular diseases (NMDs). The mRNA-based BNT162b2 and inactivated whole-virus CoronaVac are two novel COVID-19 vaccines widely used across the world that confer immune protection to healthy individuals. However, hesitancy towards COVID-19 vaccination was common for patients with NMDs early in the pandemic due to the paucity of data on the safety and efficacy in this specific patient population. Therefore, we examined the underlying factors associated with vaccine hesitancy across time for these patients and included the assessment of the reactogenicity and immunogenicity of these two vaccines.

**Methods:** Pediatric patients were screened from our NMD registry. For the vaccine hesitancy arm, those aged 8-18 years with no cognitive delay were invited to complete surveys in January and April 2022. For the reactogenicity and immunogenicity arm, patients aged 2-21 years were enrolled for COVID-19 vaccination between June 2021 to April 2022. Participants recorded adverse reactions (ARs) for 7 days after vaccination. Peripheral blood was obtained before BNT162b2 or CoronaVac and within 49 days after vaccination to measure their serological antibody responses as compared to healthy children and adolescents.

**Results:** Forty-one patients completed vaccine hesitancy surveys for both timepoints, and 22 joined our reactogenicity and immunogenicity arm of the study. Two or more family members vaccinated against COVID-19 was positively associated with intention of vaccination (odds ratio 11.7, 95% CI 1.81-75.1, *p*=0.010). Pain at the injection site, fatigue and myalgia were the commonest ARs. Most ARs were mild (75.5%, n=71/94). All 19 patients seroconverted against the wildtype SARS-CoV-2 after two doses of BNT162b2 or CoronaVac, although there was lower neutralization against the Omicron BA.1 variant.

**Discussion:** This study demonstrated vaccine hesitancy amongst patients with NMDs was influenced by family members and changed across time. BNT162b2 and CoronaVac were safe and immunogenic even for patients on low-dose corticosteroids. Future research is required to assess the durability of the COVID-19 vaccines, the effectiveness of booster doses and other routes of administration against emerging SARS-CoV-2 variants for these patients.

## 1 Introduction

Since the onset of the COVID-19 pandemic, infection by the SARS-CoV-2 virus has been associated with significant morbidity, mortality and negative socioeconomic impact throughout the world.(1-3) Certain patient populations, such as those with neuromuscular disorders (NMDs), have greater risks of severe disease and death from infections due to their muscle weakness of the chest wall or diaphragmatic muscle, cardiac involvement and immunosuppressed state.(4) Vaccination is highly effective against symptomatic infection, hospital admission and severe COVID-19 in healthy adults and children.(5-10) As such, the mRNA-based BNT162b2 and inactivated whole-virus CoronaVac vaccines are amongst the most widely used COVID-19 vaccines globally since authorization for emergency use by World Health Organization.(11-13) Based on these findings in healthy individuals, several national neurology associations and neuromuscular disease networks recommend COVID-19 vaccination for those with NMDs, but data on this important high-risk patient population specifically remain scarce.(4)

Although COVID-19 vaccination is expected to reduce infectious disease severity in the NMD population, vaccine hesitancy appears to be a major potential barrier.(14) As example, only 69.0% of the parents would want their NMD child vaccinated during the early pandemic period in December 2020.(15) It is also concerning that as little as 42.6% of those with Duchenne muscular dystrophy (DMD) were vaccinated against COVID-19 by November 2021 in Poland.(16) Their reasons for not opting for vaccination during this early, pre-Omicron variant period included the potential for reduced efficacy due to their use of immunosuppressive or immunomodulating therapies and uncertainties regarding possible interactions between the vaccines and treatments for NMDs.(17-19) Despite the availability of two different COVID-19 vaccine types, BNT162b2 and CoronaVac, in our locality, our NMD patients also appeared reluctant on vaccination. Some of these patients had cited the risks of adverse effects, disease complications and reduced efficacy as some of their main concerns.

The reason that there has been a paucity of NMD-specific safety and immunogenicity data despite the rollout of the many types of COVID-19 vaccines is because NMD is a group of rare diseases and NMD patients are hesitant to volunteer for receiving novel vaccines, and so large-scale studies had not yet been possible. Therefore, scientific evidence on COVID-19 vaccination in NMD patients have been based on small cohorts only thus far. For 14 adult NMD patients, BNT162b2 and mRNA-1273 vaccines seem safe and immunogenic.(20) The mRNA-1273 vaccine achieved robust humoral and cellular immune responses in 100 adult patients with myasthenia gravis.(21) Unfortunately, there are still no available safety and immunogenicity data on COVID-19 vaccines in children with NMDs. Importantly, there has been no previous research on the inactivated COVID-19 vaccines in adult or pediatric patients with NMDs, including immunogenicity against the novel variants, such as Omicron.

Therefore, this study investigated in-depth the underlying reasons and temporal changes in vaccine hesitancy for pediatric patients with NMDs during the Omicron wave in 2022. Additionally, we assessed the safety and immunogenicity of BNT162b2 and CoronaVac by recording adverse effects and measuring serum antibody levels and neutralization against the wild type (WT) SARS-CoV-2 virus and Omicron B.1.1.529 variant.

## 2 Materials and methods

### 2.1 Study Population

All participants were screened from the Hong Kong (HK) NMD registry.(22) This registry has been approved by the Institutional Review Board (IRB) and collects patient and clinician-reported demographic and clinical information from those with confirmed diagnoses of NMDs after their consent.(22) The diagnosis was determined by clinical pediatric neurologists and supported by genetic testing and/or muscle biopsy results. The neurology clinical study team is based at HK Children’s Hospital, Duchess of Kent Children’s Hospital at Sandy Bay and Queen Mary Hospital that receives referrals from throughout the entire HK territory for clinical care and research in pediatric NMDs.(22) Enrolment in the vaccine hesitancy survey arm of the study required patients to be 8-18 years old with no cognitive delay. In another arm of this study, patients aged 2-21 years were invited for COVID-19 vaccination to study the reactogenicity and immunogenicity of the BNT162b2 and CoronaVac vaccines. Potential participants needed to be in stable condition. A patient could join either or both arms of the study, if eligible (Supplementary Figure 1).

### 2.2 COVID-19 Vaccine Hesitancy Survey

Patients received and completed the first survey through online or phone interviews in January 2022. The survey was based on our previous publication on 2,609 healthy adolescents, supplemented with more tailored queries pertinent to NMD patients and younger age groups that were added into this study.(23) In summary, it included 21 yes/no or multiple-choice questions on patient demographics, presence of medical complexity, history of past COVID-19 infection, influenza and COVID-19 vaccination, intention of receiving COVID-19 vaccination and the reasons for their choice (Supplementary file). Concerns about receiving COVID-19 vaccination included perception of risks, challenges in access to vaccination centers, adverse effects, less efficacy than their healthy counterparts and vaccine-drug interactions that potentially affect their current NMD treatments. The expected time required to complete the survey was 15 minutes. A follow-up survey was sent to patients in April 2022 to longitudinally assess changes in attitudes, hesitancy and associated reasons shortly after the peak of the first major COVID-19 wave due to the SARS-CoV-2 Omicron variant in HK.

### 2.3 Reactogenicity and Immunogenicity Study of COVID-19 Vaccines

The reactogenicity and immunogenicity arm is a sub-study under the registered Coronavirus disease-19 (COVID-19) Vaccination in Adolescents and Children (COVAC) (NCT04800133). The COVID-19 vaccines were administered at the Community Vaccination Centers research sites supported by the University of Hong Kong (HKU) and the HK Government’s COVID-19 Vaccination Program. Patients received 2 doses of either BNT162b2 or CoronaVac, given 21 or 28 days apart, respectively, followed by an option of either vaccine types as a third dose at least 28 days after their second dose. The dosages of BNT162b2 were 0.3 mL (equivalent to 30 μg of COVID-19 mRNA vaccine embedded in lipid nanoparticles) for participants aged ≥12 years. One-third fractional dose of BNT162b2 was administered to children aged 5-11 years. The dosage of CoronaVac was 0.5 mL (600 SU, equivalent to 3 μg, of inactivated SARS-CoV-2 virus as antigen) for all ages.

Vaccine recipients were monitored for 30 mins after each injection and reported the types, duration, and severity of adverse reactions (ARs) in a diary by an online or paper format for 7 days after vaccination. Peripheral blood was obtained before the first dose, second dose, 7-43 days after the second dose and 14-49 days after the third dose (if any) for measuring the serological antibody responses. The SARS-CoV-2 S receptor-binding domain (S-RBD) IgG enzyme linked immunosorbent assay (ELISA) (Chondrex Inc, Redmond, USA) and surrogate virus neutralization test (sVNT) (GenScript, New Jersey, USA) performed in our laboratory on the serum isolated from blood samples of patients had been validated and described in our previous publication.(24) Levels of S-RBD IgG are expressed as optical density (OD_450_). The cut-off considered as seroconversion was OD_450_ ≥0.50, while values below would be inputted as 0.25.(25) Neutralizing antibodies against SARS-CoV-2 WT and Omicron BA.1 were evaluated by sVNT with inhibition percentages (%) as the readout.(26) The cut-off for positive neutralizing antibody inhibition was ≥30%, and values below 30% would be inputted as 10%. Data from healthy children and adolescents were retrieved from our previous publication and COVAC study for comparison to this NMD cohort.(27)

Social and contact avoidance was common during this study period when the HK Omicron wave occurred, particularly for vulnerable patients as neurological and respiratory complications surged rapidly.(28) Therefore, some NMD patients were only able to attend our vaccination research sites and provide blood samples at the time-point of 7-43 days after the second dose.

### 2.4 Statistical Analysis

Associations between the categorical variables (presence of medically complexity, history of past COVID-19 infection, influenza and COVID-19 vaccination) and intention of receiving COVID-19 vaccination (i.e., have received or plan to receive *vs* do not plan to receive COVID-19 vaccination) were analyzed by the Fisher’s exact test. Intention of receiving COVID-19 vaccination between January and April 2022 was compared by the Cochran’s Q test. Additionally, reasons for receiving the COVID-19 vaccines in January and April 2022 were compared by the Fisher’s exact test. S-RBD IgG levels and sVNT % inhibition against WT were compared between NMD patients and the healthy population, also between the two vaccine types, by the Mann-Whitney U test. Comparisons between the sVNT% inhibition level against WT and Omicron BA.1 for each patient were computed using the Wilcoxon matched-pairs signed-rank test. *p*<0.05 was considered statistically significant. Data analyses and graphing were performed using GraphPad Prism (version 9.3.1).

### 2.5 Standard Protocol Approval, Registration and Patient Consent

The NMD patient registry and COVAC were approved by the HKU/Hospital Authority HK West Cluster IRB Committee (UW19-356 and UW 21-157, respectively). Written informed consent was obtained from adult participants or legally authorized representatives of the child participants.

## 3 Results

Of the 136 patients in the HK NMD registry, 52 were sent invitations to complete the COVID-19 vaccine hesitancy survey, while the others did not fulfill criteria due to age or cognitive delay. Most patients, which were 41 (78.8%) of the 52, completed both the first and follow-up surveys. Eighteen (43.9%) of 41 who filled the survey had spinal muscular atrophy (SMA), while 26 (63.4%) had complex medical needs, including wheelchair mobility, tube or gastrostomy tube feeding, ventilator use, or brace or spinal surgery for scoliosis (Supplementary Table 1).

Two or more family members vaccinated against COVID-19 was positively associated with a higher intention of vaccination (OR: 11.7, 95% CI: 1.81-75.1, p=0.010) (Table 1). Patients who received an influenza vaccine in the last three consecutive years tended to have higher intention of receiving COVID-19 vaccines, albeit not reaching statistical significance (24 of 30 vs 5 of 11, or 80.0% vs 45.5%, p=0.052). The major reasons that the NMD patients favored COVID-19 vaccination included their hopes for preventing infection (26 of 40, or 65.0%), protecting their family (16, or 40.0%) and returning to normal life (14, or 35.0%) (Table 2). Their concerns regarding COVID-19 vaccination included adverse effects that could be potentially worse than the healthy population (9 of 41, 22.0%), safety (9, or 22.0%), suitability (6, or 14.6%), effects on current NMD treatments (4, or 9.8%) and reduced efficacy (4, or 9.8%) (Figure 1). There were also 5 (12.2%) of 41 patients who expressed that their intention of vaccination depended on the progress of the pandemic. Indeed, in April 2022, which was shortly after the peak of the Omicron wave in HK, more respondents had or planned to receive the COVID-19 vaccines than in January 2022 (97.6% vs 73.3%, p=0.003) (Supplementary Table 2). Also, 30 (73.2%) patients expressed their intention of receiving future boosters, if necessary, in April 2022 (Supplementary Table 2). Twenty (48.8%) of 41 patients expressed a vaccination history/intention of vaccination for at least 1 dose of BNT162b2 (B) or CoronaVac (C) in April 2022 (Supplementary Table 3).

**Table 1:**
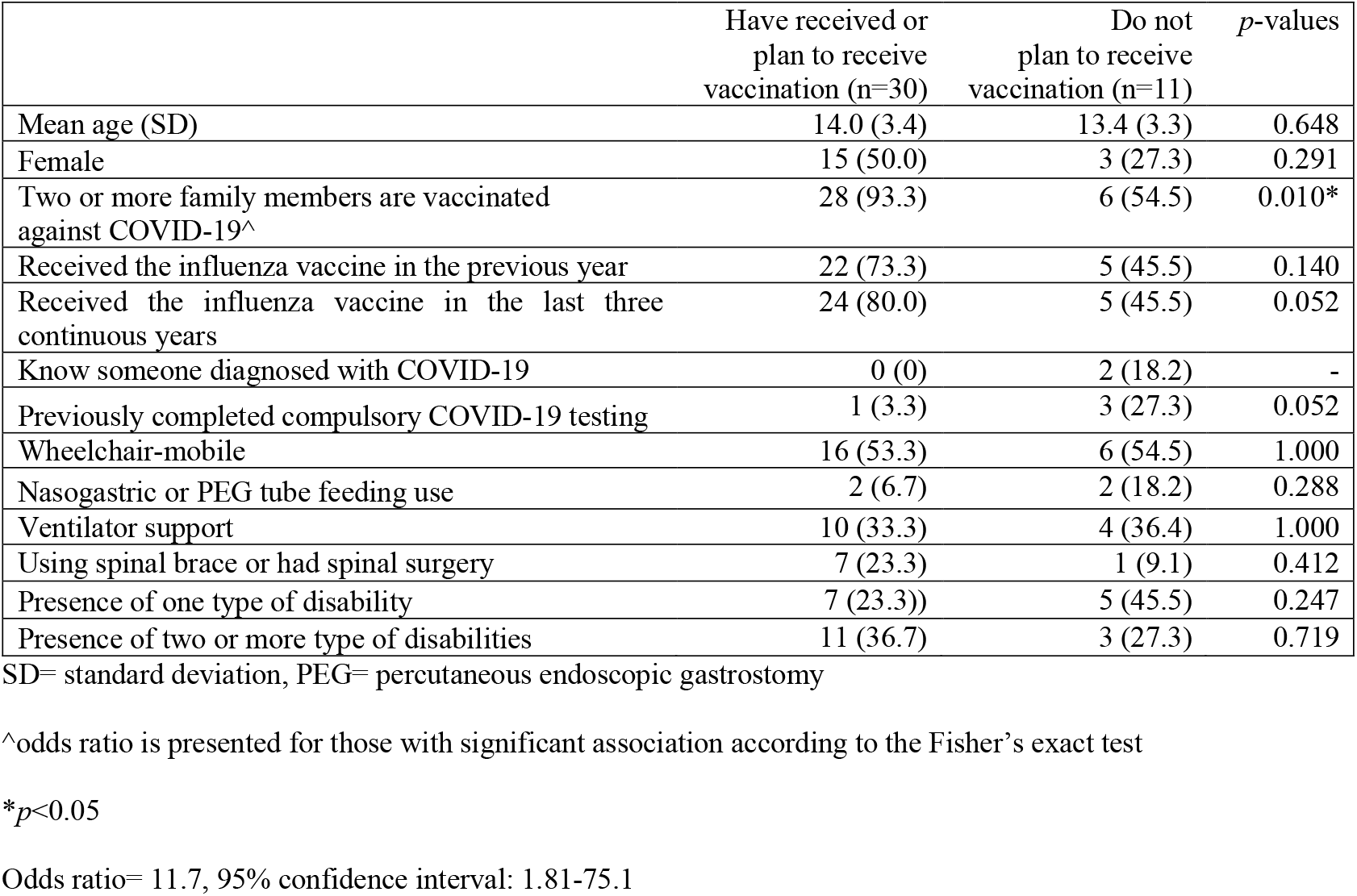
Factors associated with intention of receiving COVID-19 vaccination in patients with neuromuscular diseases in January 2022.

**Table 2:** Major reasons for receiving COVID-19 vaccination in patients with neuromuscular diseases

|  | January 2022 | April 2022 |  |
| --- | --- | --- | --- |
|  | No. (%) | No. (%) | *p*-values |
| Planning to vaccinate/ already vaccinated | n=30 | n=40 |  |
| Worried to be infected | 17 (56.7) | 26 (65.0) | 0.620 |
| To return full-scale face-to-face teaching | 16 (53.3) | 16 (40.0) | 0.335 |
| To protect their family | 17 (56.7) | 14 (35.0) | 0.091 |
| To return to normal life | 19 (63.3) | 13 (32.5) | 0.015* |
| I think I am high risk group of COVID-19 | 7 (23.3) | 12 (30.0) | 0.596 |
| Because people around me received vaccination | 5 (16.7) | 11 (27.5) | 0.391 |
| I am tired of the social distancing policies | 9 (30.0) | 10 (25.0) | 0.787 |
| I want to socialize with large groups of friends | 3 (10.0) | 10 (25.0) | 0.132 |
| I want to socialize with participate in large-scale events | 2 (6.7) | 9 (22.5) | 0.100 |
| I want to travel | 14 (46.7) | 8 (20.0) | 0.022* |
| I want to play sports at school with friends or in a competition without wearing mask | 11 (36.7) | 7 (17.5) | 0.098 |
| To play music safely<br>E.g.: wind instruments, band, choir | 6 (20.0) | 7 (17.5) | 1.000 |
| I want to join study tours<br>E.g.: day trip, school visits or other extra-curricular activities | 12 (40.0) | 5 (12.5) | 0.011* |

\* $p < 0.05$

**Figure 1:**
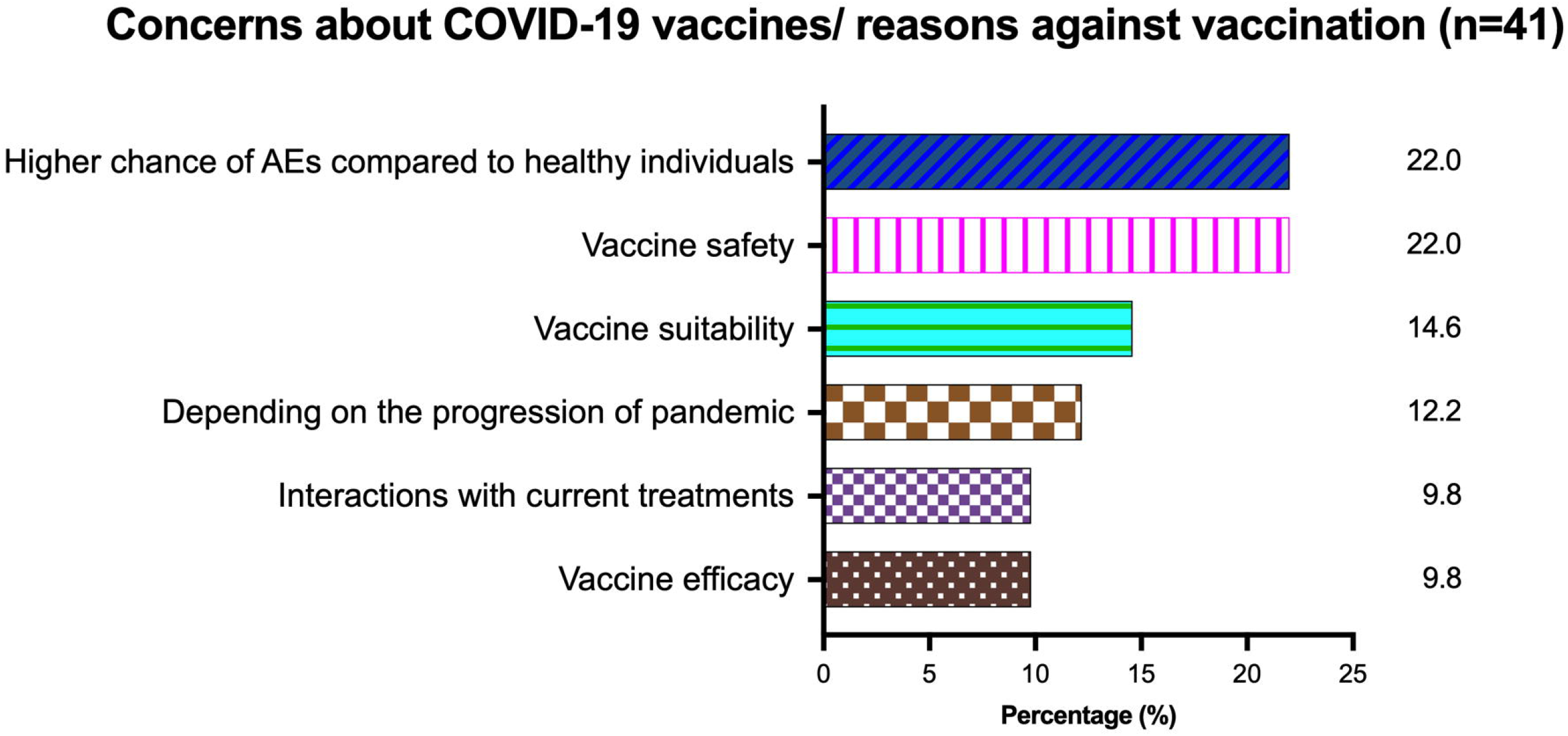
Major concerns about receiving COVID-19 vaccination in patients with neuromuscular diseases, AEs=adverse effects. The three commonest concerns from patients with neuromuscular diseases were having a higher chance of AEs compared to healthy individuals (22.0%), safety (22.0%) and suitability (14.3%).

Forty-eight patients were invited from the NMD patient registry to complete the reactogenicity and immunogenicity arm from June 2021 to April 2022, while the others were not recruited due to the age exclusion criterion. Twenty-two (45.8%) patients joined the study (Supplementary Table 4). Nine patients had DMD, 7 patients had SMA, 3 patients had congenital myopathy (CM) and 1 patient had Becker muscular dystrophy, chronic inflammatory demyelinating polyneuropathy (CIDP) or myotonic disorder. Fourteen (63.6%) were in the late ambulatory or wheelchair mobile stage. All 9 DMD patients were on corticosteroids (ranges of dosage: 10-30 mg/day or 0.30-0.74 mg/kg/day). Seven SMA patients were on nusinersen or risdiplam. One CIDP patient was on intravenous immunoglobulin therapy infused regularly (dosage: 2 g/kg/3months). There were 16 (72.7%) and 6 (27.3%) of 22 patients who received two doses of BNT162b2 (BB) or CoronaVac (CC), respectively. One case of CM and DMD each had COVID-19 before enrollment into the study, while 1 patient with SMA reported contracting COVID-19 3 weeks after the first dose, and all three patients fully recovered subsequently.

Pain at the injection site (BB: 3 of 6, 50.0%, 6 of 11, CC: 54.5%), fatigue (BB: 4 of 6, 66.7%, 5 of 11, CC: 45.5%) and myalgia (BB: 1 of 6, 16.7%, 3 of 11, CC: 27.3%) were the commonest ARs. Most ARs were mild (75.5%, n=71/94). No severe adverse events, such as apparent NMD deterioration, hospitalization, life-threatening complications, disabilities or deaths occurred (Figure 2).

**Figure 2:**
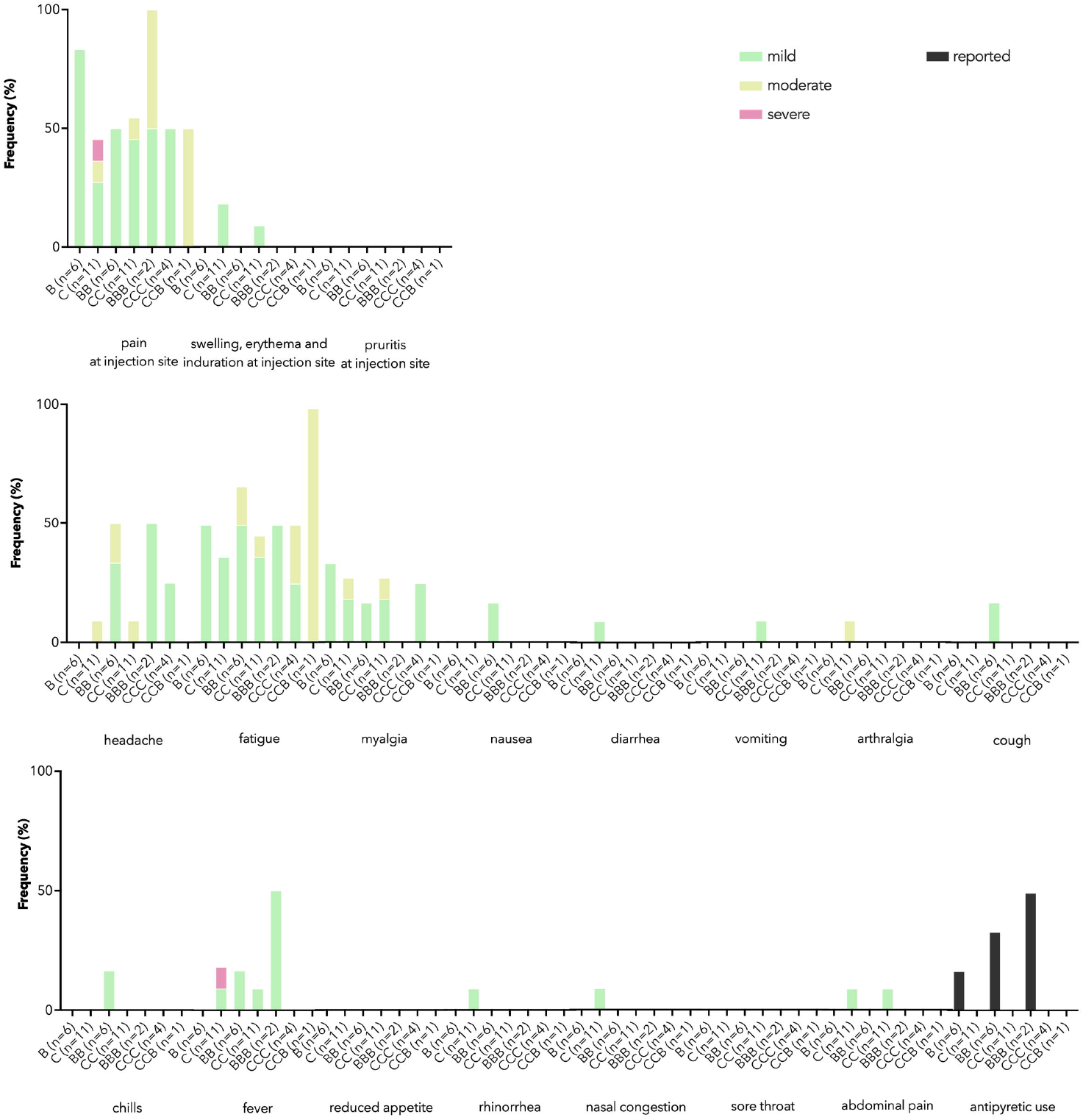
Reactogenicity for patients with neuromuscular diseases Pain at injection site, fatigue and myalgia were the three commonest adverse reactions (ARs). Most ARs were mild. No patient reported any of the following symptoms within 7 days after vaccination: pruritis at injection site, reduced appetite or sore throat. There were no vaccine-related severe adverse events, such as hospitalizations, life-threatening complications, disabilities or deaths. B, 1 dose of BNT162b2; BB, 2 doses of BNT162b2; C, 1 dose of CoronaVac; CC, 2 doses of CoronaVac; BBB, 3 dose of BNT162b2; CCC, 3 doses of CoronaVac; CCB, 2 doses of CoronaVac and 1 dose of BNT162b2.

All 19 patients with NMDs seroconverted against WT after BB or CC (Figure 3a). NMD patients had similar antibody responses compared to healthy children and adolescents (ELISA-CC: 2.03 vs 1.59; ELISA-BB : 2.19 vs 2.86; sVNT-CC: 85.2% vs 83.2%) (Figure 3a; Figure 3b). Although after receiving CC, the group on corticosteroids had lower S-RBD IgG than those not on corticosteroids (OD450: 1.82 vs 2.37, p=0.02), corticosteroids did not affect the seroconversion rate (Supplementary Table 5). There was lower neutralization against Omicron BA.1. The median sVNT inhibition against WT and Omicron BA.1 was 96.3% and 10.0% after BB (p=0.063) (Figure 3c), respectively, while it was 85.2% and 10.0% after CC (p<0.001) (Figure 3d).

**Figure 3:**
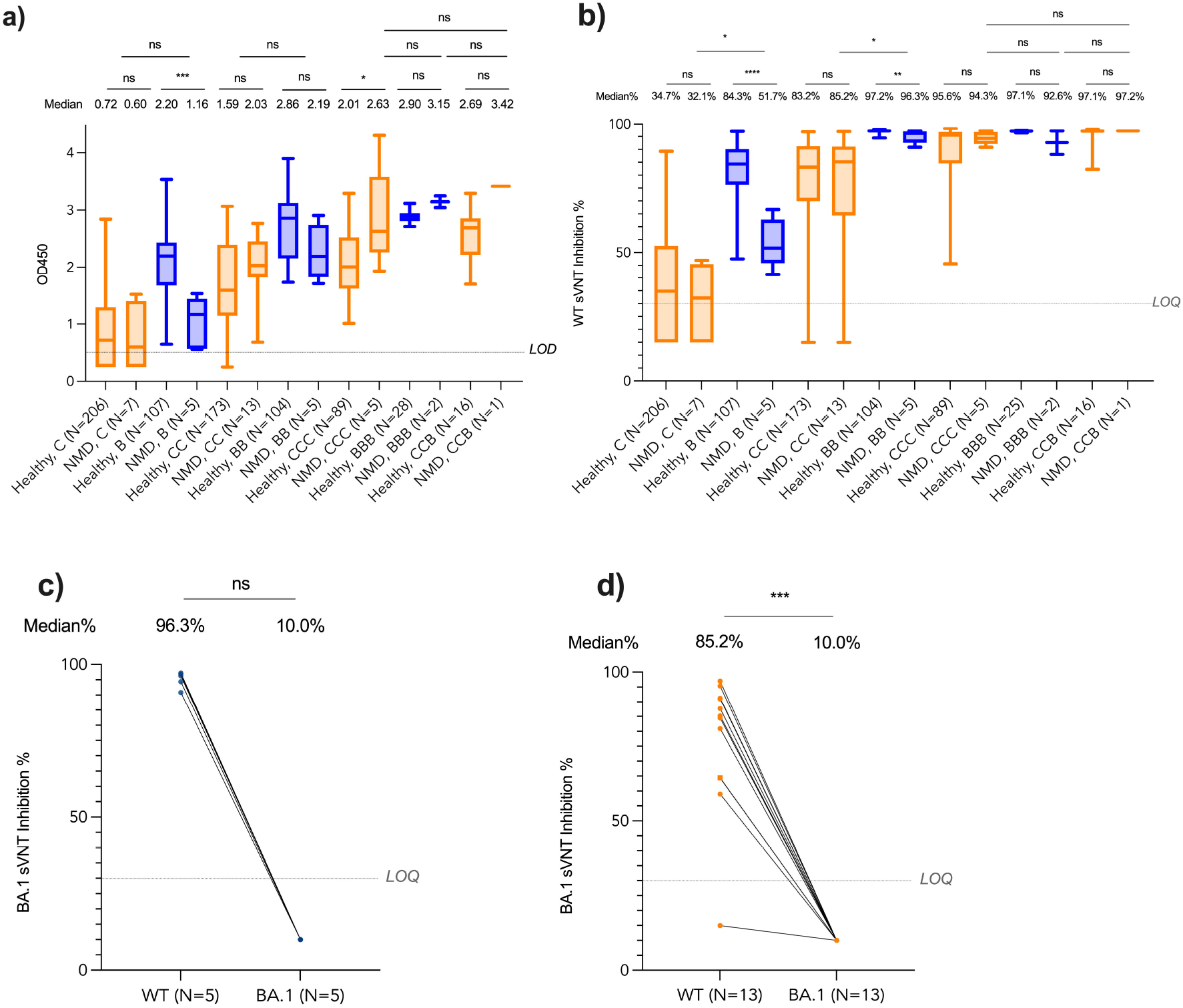
Immunogenicity of BNT162b2 and CoronaVac for patients with neuromuscular diseases Antibody responses were determined before the first dose, second dose, 7-43 days after the second dose and 14-49 days after the third dose of BNT162b2 or CoronaVac in patients with neuromuscular diseases (NMDs) (n=19). Three patients who had COVID-19 infection were excluded from the final analysis. Immunogenicity data from healthy children and adolescents used for comparisons with patients with NMDs were retrieved from our previous publication and COVAC study.(27) (a) and (b) All patients with NMDs successfully seroconverted by enzyme linked immunosorbent assay (ELISA) against wildtype SARS-CoV-2 virus (WT) after receiving two doses of BNT162b2 and CoronaVac. These patients showed similar antibody responses when compared to the healthy children and adolescents. (c) and (d) surrogate virus neutralization test (sVNT) after BNT162b2 and CoronaVac was significantly lower against the SARS-CoV-2 Omicron BA.1 variant compared to WT. (c) The median % of inhibition level in the BB group was lower at 10.0% for Omicron BA.1 than WT at 96.3% (p>0.05). (d) The median % of inhibition level in the CC group was lower at 10.0% for Omicron BA.1 than WT at 85.2% (p<0.001). B, 1 dose of BNT162b2; BB, 2 doses of BNT162b2; C, 1 dose of CoronaVac; CC, 2 doses of CoronaVac; BBB, 3 dose of BNT162b2; CCC, 3 doses of CoronaVac; CCB, 2 doses of CoronaVac and 1 dose of BNT162b2. BNT162b2 was represented as blue while CoronaVac was represented as orange color. Patients with NMDs using corticosteroids (deflazacort or prednisolone) daily were indicated as square dots in (c) & (d). *p<0.05, **p<0.01, ***p<0.001, ****p<0.0001) or NS (not significant).

## 4 Discussion

This study demonstrated COVID-19 vaccination in family members is highly influential on the intention of receiving the vaccines for pediatric patients with NMDs, and those who received either the mRNA-based or inactivated whole-virus vaccines did not encounter severe ARs and had antibody responses similar to their healthy counterparts. This is consistent with the notion that family decision and support are key factors on COVID-19 vaccination for pediatric populations, as several recent studies observed this finding for healthy adolescents and children with neurodevelopmental disorders. (23, 29, 30) Additionally, patients who received the influenza vaccines in the recent consecutive years tended towards having the greater intention of COVID-19 vaccination, and we speculate this was due to higher vaccine confidence and complacency.(14, 31) These patients with NMDs were highly concerned about becoming infected, which was their main underlying reason for receiving the COVID-19 vaccines. This information will be useful for patient counseling as they continued to raise questions in our clinic regarding the need for the third dose and subsequent boosters in the future.

This is the first study to investigate the safety of the novel COVID-19 vaccines in children with NMDs, which showed BNT162b2 and CoronaVac were well tolerated. There were similar profiles of ARs between pediatric patients with NMDs, our healthy cohort, as well as adolescents and adults with NMDs and multiple sclerosis who received two doses of BNT162b2 or mRNA-1273.(20, 27, 32, 33) This is also the first study to demonstrate the inactivated whole-virus COVID-19 vaccine, CoronaVac, is immunogenic in pediatric patients with NMDs. Antibody responses were robust, an observation which was consistent with adolescent and adult patients with NMDs and myasthenia gravis who were able to generate antibody responses against WT.(20, 21, 33) It is reassuring that even for pediatric patients with NMDs and on corticosteroids, all patients had successful seroconversion after at least two doses of BNT162b2 or CoronaVac, which was also observed in adolescent and adult patients who received BNT162b2.(20, 33) Additionally, there were no apparent interactions between the COVID-19 vaccines and treatments for NMDs, and our cohort of patients did not encounter NMD-related complications or hospitalization.

Importantly, our study included immunogenicity data against Omicron, which was first detected in southern Africa and became the dominant strain that causes SARS-COV-2 infections since late 2021.(34) Different studies showed reduced vaccine effectiveness (VE) of BNT162b2 and CoronaVac against infection or mild COVID-19 due to the Omicron variant, including children and adolescents.(7, 8, 35, 36) Our findings revealed reduced neutralizing activity against Omicron BA.1 in pediatric patients with NMDs. As neutralization correlates with protective efficacy against symptomatic COVID-19, we expect breakthrough infections would be more common in NMD patients due to Omicron than pre-Omicron variants, similarly with the rest of the population. A recent study in HK indicated that the VE of BNT162b2 and CoronaVac against infection would be 55.0% after two doses of vaccination in adolescents.(37, 38) On the other hand, studies showed that T cell responses in vaccinated adults were preserved against the BA.1 variants,(39, 40) which is correlated with clinical protection against severe diseases.(41) Therefore, patients with NMDs should become vaccinated to attain protection against severe COVID-19. Further boosters may enhance neutralization responses against Omicron subvariants for maximal protection.

There were several limitations in this study. First, it was not possible for participants to be enrolled into a study with a blinded and randomized design because these patients are already hesitant to receive novel vaccines and restriction on their choice on the type of vaccine would be an additional deterrent. There can be potential selection bias because more older males favored BNT162b2. However, the current findings have more real-life applicability and reflect the reality of outcomes for patients who can choose between vaccines. Reactogenicity and immunogenicity results were not available for all the timepoints, as we encouraged patients to receive the vaccines as soon and conveniently as possible for protection from severe COVID-19. This is because informed choice and prompt preventative treatment for overcoming a surging wave of infection-related deaths in high-risk patients during the peak of our pandemic period is paramount and should be respected. Finally, the small sample size limits the generalizability of the study conclusions, but these are directly applicable to an important group of patients with a rare disease and are at high risk of severe infection-related complications. These results greatly contribute to the currently available scientific evidence that serves as the basis for appropriate clinical practice recommendations and policy-making decisions for patients with NMDs.

Taken together, these present findings and overall evidence support the routine schedule of vaccination for patients with NMDs, and the dosages of immunosuppressives used for the treatment of NMDs in relation to the BNT162b2 and CoronaVac vaccines are inconsequential.(4) We recommend that counseling for these patients should incorporate these informative points and to include several close relatives, if possible, because their decisions appear to be strongly influential towards vaccine hesitancy. Reassurance by reminding the patients about their tolerance to other vaccines, such as influenza, can also be considered. Future research is required to confirm that these counseling techniques are effective on addressing vaccine hesitancy. Additionally, as high as 26.8% (11 of 41) of our patients who completed the primary series have already expressed reluctance towards boosters in April 2022. Questions remain in terms of the safety, efficacy, long-lasting T cell immunity and durability of booster doses against other emerging variants,(42) such as Omicron BA2.75, BA.5, XBB, BQ1.1 and BF.7 for these patients. Finally, innovative aerosolized vaccines are under development that aim to induce mucosal immunity and thereby reduce viral transmission.(43) This method of inhibiting the initial step of infection offers a pathway towards ending the pandemic, which would be most promising if such route of immunization can be achieved for as many individuals as possible. Whether these aerosolized vaccines are effective for NMD patients, who have impaired respiratory function and inspiratory lung volumes, is intriguing and should be studied further.

## Data Availability

All data produced in the present work are contained in the manuscript

## 7 Conflict of Interest

All authors declare that there is no conflict of interest.

## 8 Author Contributions

Michael Kwan Leung Yu was involved in the recruitment, performing the statistical analysis, interpretation of results and drafting this manuscript.

Sophelia Hoi Shan Chan was involved in the recruitment, clinical care, interpretation of results and drafting this manuscript.

Samuel Cheng was involved in conducting the ELISA and sVNT assay as well as the interpretation of results.

Daniel Leung was involved in the recruitment, study design, interpretation of results and drafting this manuscript.

Sau Man Chan was involved in the recruitment, study design and clinical care. Amy Suen Ka Yan was involved in the study administrative procedures.

Wilfred Hing Sang Wong was involved in supervising the statistical analysis.

Malik Peiris was involved in supervising the ELISA and sVNT assay as well as the interpretation of results.

Yu-Lung Lau was the principal investigator of this project and was involved in the recruitment, study design, clinical care, interpretation of results and drafting this manuscript.

Jaime S Rosa Duque was involved in the recruitment, study design, clinical care, interpretation of results and drafting this manuscript.

## 9 Funding

This study was funded by the Health Bureau of the Government of Hong Kong, COVID19F02, COVID19F10 and COVID19F12.

## 10 Acknowledgements

This work was supported by the research grant COVID19F02, COVID19F10, and COVID19F12 from the Health Bureau of the Government of Hong Kong. We thank Dr. Davy Chun-Wai Lee and Mr. Koon-Wing Chan at the Department of Paediatrics and Adolescent Medicine, LKS Faculty of Medicine, HKU for their laboratory management of the collected blood samples.

## 12 Data Availability Statement

The original contributions presented in the study are included in the article/supplementary material, further inquiries can be directed to the corresponding author/s.

---

**Supplementary Table 1:**
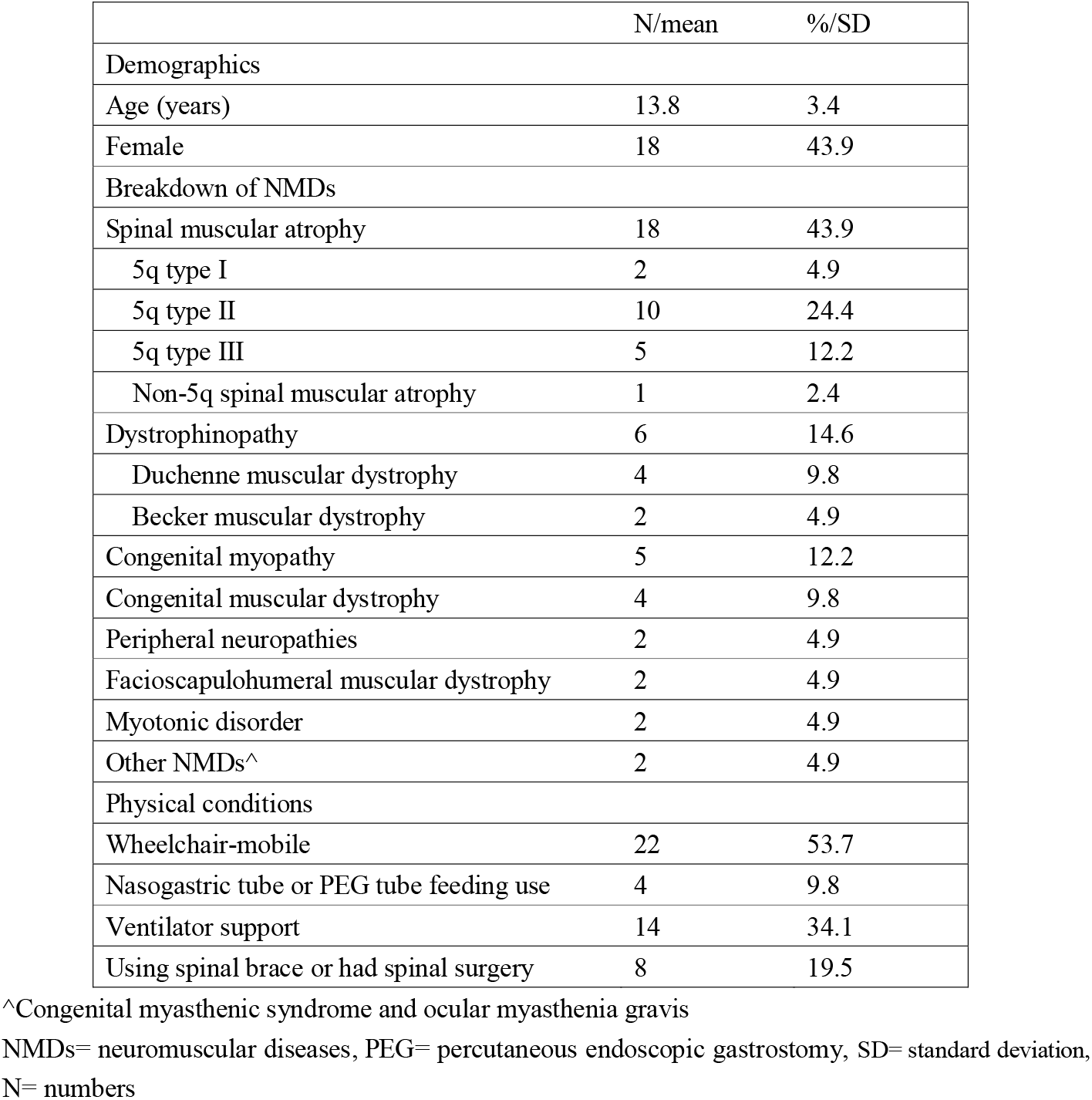
Demographic and clinical characteristics of the patients with neuromuscular diseases in COVID-19 vaccine hesitancy survey arm of the study

**Supplementary Table 2:**
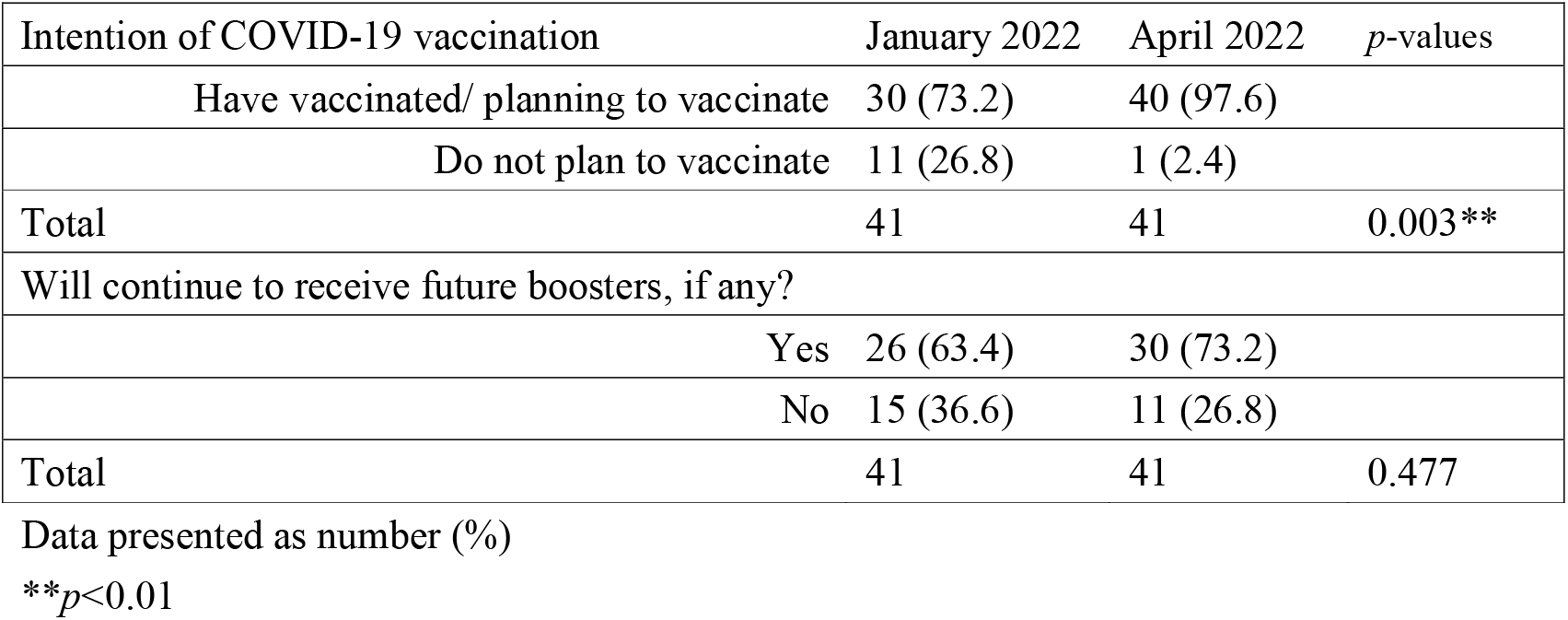
Intention of COVID-19 vaccination in patients with neuromuscular diseases in January and April 2022

**Supplementary Table 3:**
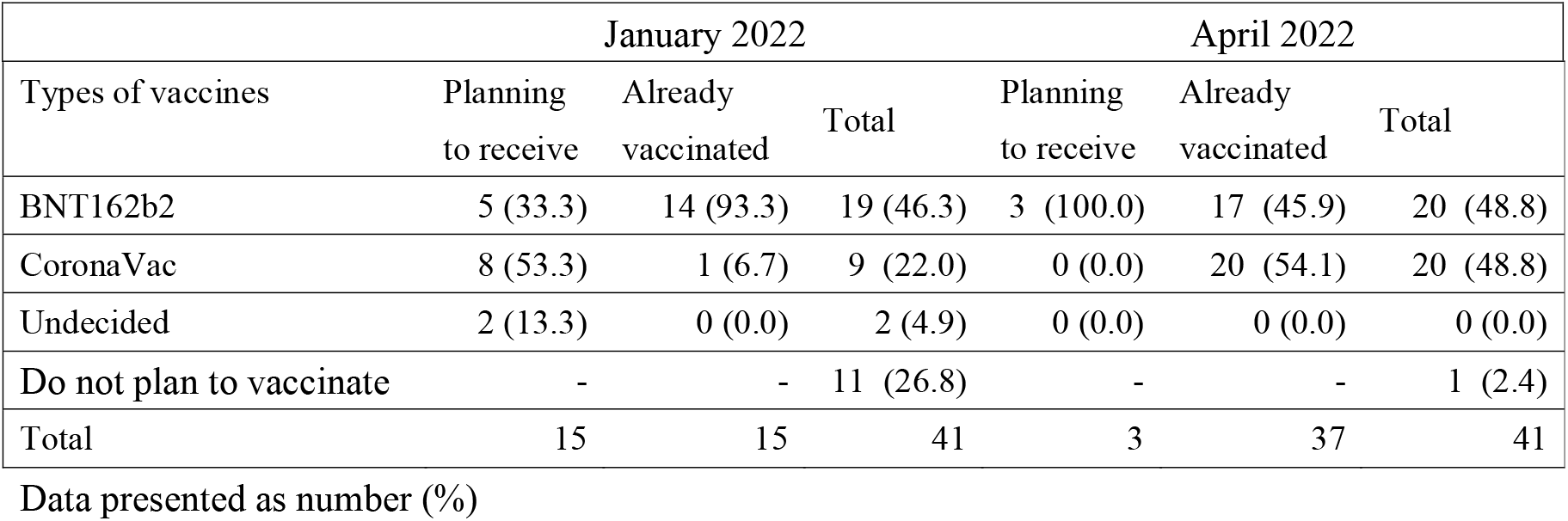
Preference on COVID-19 vaccines in patients with neuromuscular diseases

**Supplementary Table 4:**
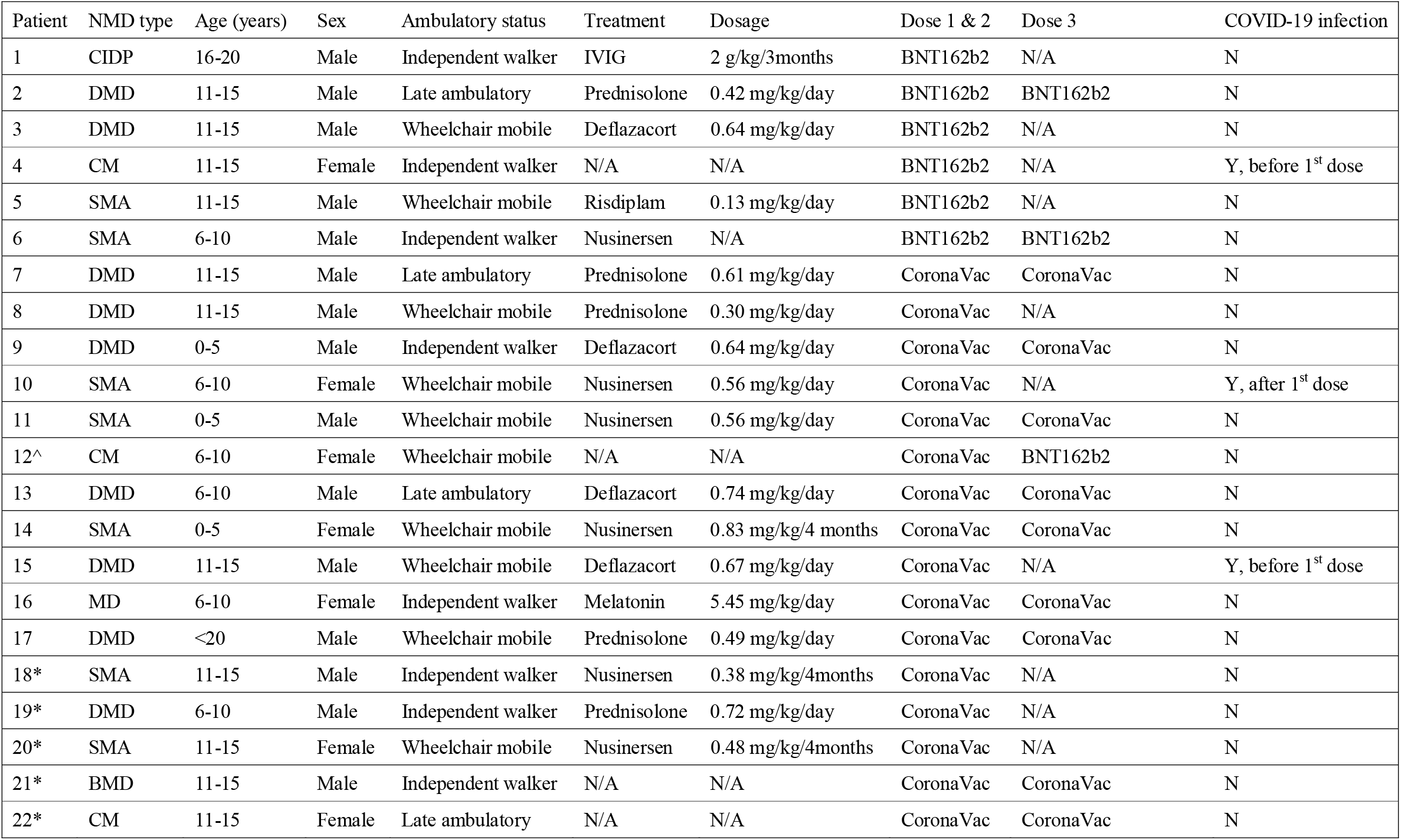
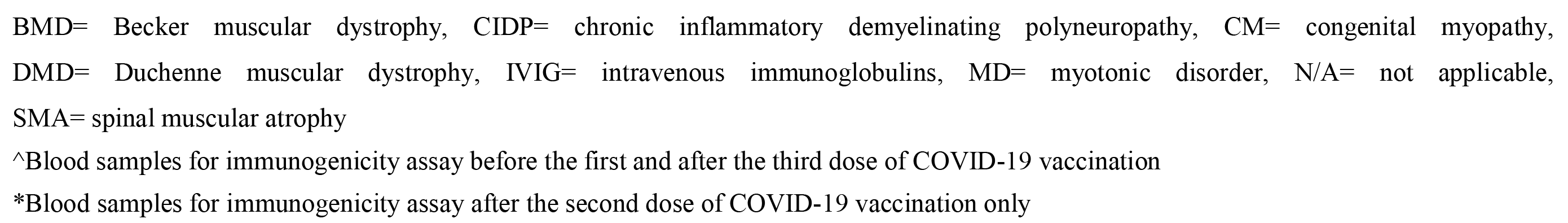
Demographic and clinical characteristics of the patients with neuromuscular diseases in the COVID-19 vaccine reactogenicity and immunogenicity arm of the study

**Supplementary Table 5:**
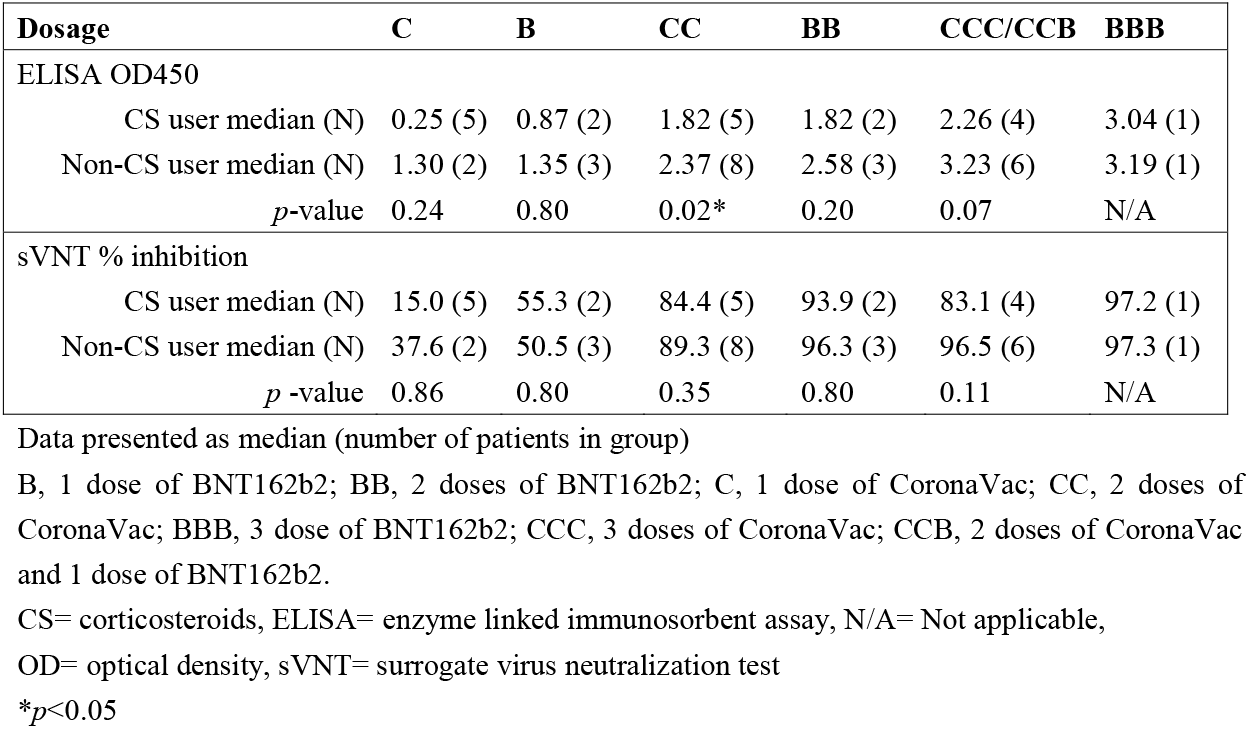
Comparison of immunogenicity for patients with neuromuscular diseases by corticosteroids use

| **Dosage** | **C** | **B** | **CC** | **BB** | **CCC/CCB** | **BBB** |
| --- | --- | --- | --- | --- | --- | --- |
| **ELISA OD450** |  |  |  |  |  |  |
| CS user median (N) | 0.25 (5) | 0.87 (2) | 1.82 (5) | 1.82 (2) | 2.26 (4) | 3.04 (1) |
| Non-CS user median (N) | 1.30 (2) | 1.35 (3) | 2.37 (8) | 2.58 (3) | 3.23 (6) | 3.19 (1) |
| *p*-value | 0.24 | 0.80 | 0.02* | 0.20 | 0.07 | N/A |
| **sVNT % inhibition** |  |  |  |  |  |  |
| CS user median (N) | 15.0 (5) | 55.3 (2) | 84.4 (5) | 93.9 (2) | 83.1 (4) | 97.2 (1) |
| Non-CS user median (N) | 37.6 (2) | 50.5 (3) | 89.3 (8) | 96.3 (3) | 96.5 (6) | 97.3 (1) |
| *p* -value | 0.86 | 0.80 | 0.35 | 0.80 | 0.11 | N/A |

Data presented as median (number of patients in group)
B, 1 dose of BNT162b2; BB, 2 doses of BNT162b2; C, 1 dose of CoronaVac; CC, 2 doses of CoronaVac; BBB, 3 dose of BNT162b2; CCC, 3 doses of CoronaVac; CCB, 2 doses of CoronaVac and 1 dose of BNT162b2.
CS= corticosteroids, ELISA= enzyme linked immunosorbent assay, N/A= Not applicable,
OD= optical density, sVNT= surrogate virus neutralization test
\* $p < 0.05$

**Supplementary Table 6:**
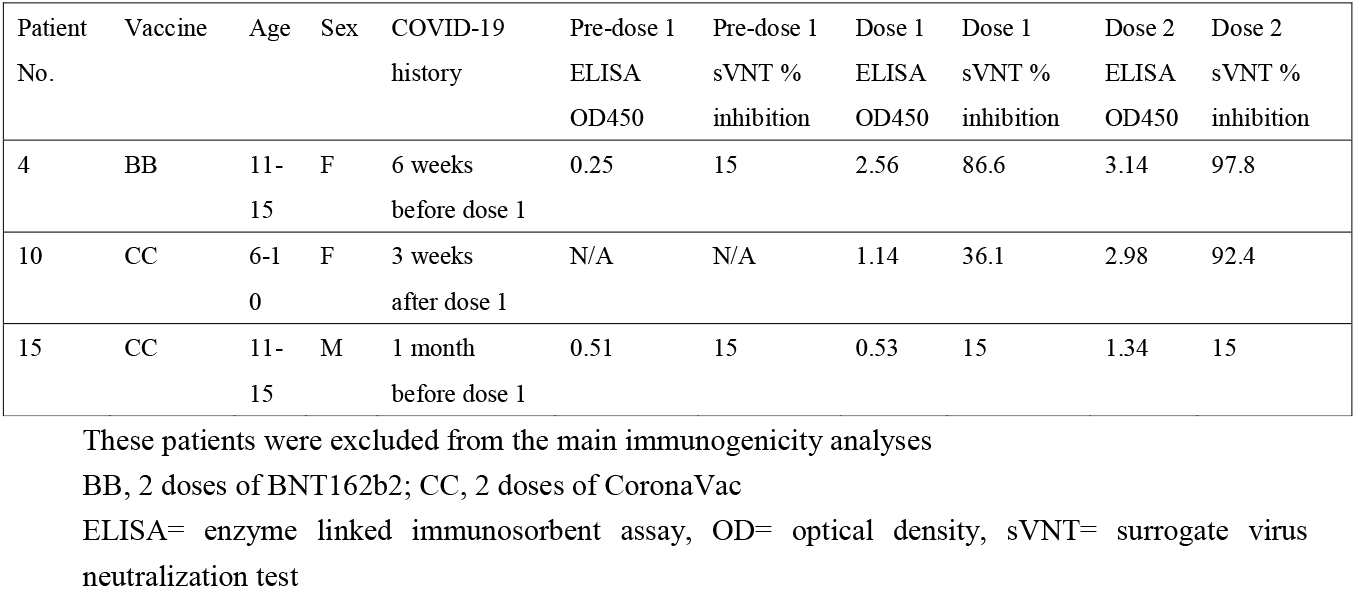
Immunogenicity of patients with neuromuscular diseases who had COVID-19 infection

**Supplementary Figure 1:**
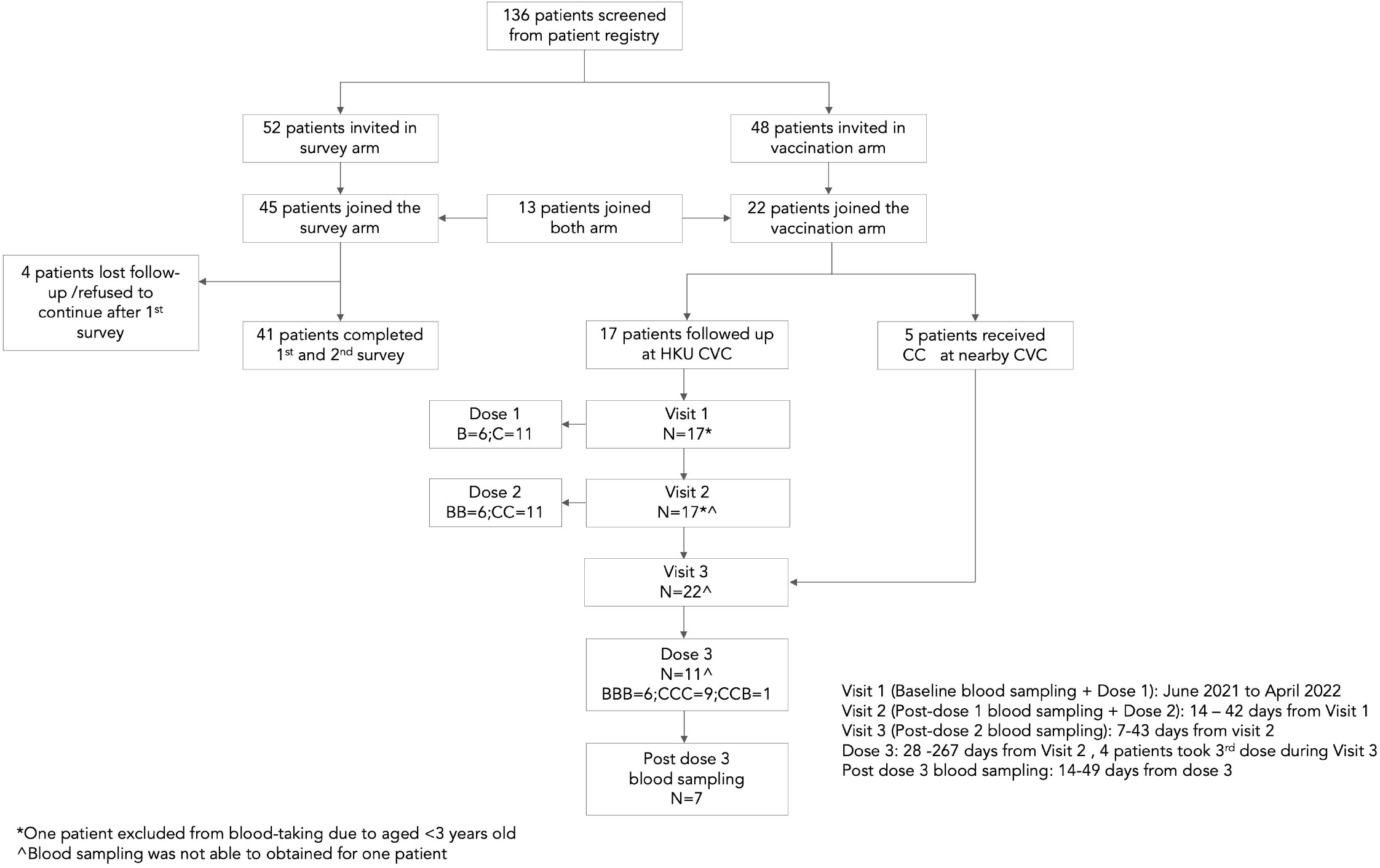
Flow diagram of study participants 136 patients were screened from the patient registry. Fifty-two patients were invited to complete the hesitancy survey arm, and 48 patients were invited to join the reactogenicity and immunogenicity arm of the study. Forty-five patients completed the first hesitancy survey, and 41 (91.1%) of them completed both first and second surveys. For the reactogenicity and immunogenicity arm of the study, 22 patients joined and 17 were inoculated with BNT162b2 or CoronaVac at our HKU Community Vaccination Centers (CVCs) research site. These patients recorded adverse reactions in a 7-day diary system after vaccination for reactogenicity/safety analyses and had blood sampling. Additionally, 5 patients who received two doses of CoronaVac at nearby CVCs and had blood sampling after the second dose.

## Notes

### Competing Interest Statement

The authors have declared no competing interest.

### Clinical Trial

NCT04800133

### Author Declarations

IRB Committee of The University of Hong Kong and Hospital Authority HK West Cluster gave ethical approval for this work

