## Supplemental Material for "Vaccine hesitancy, reactogenicity and immunogenicity of BNT162b2 and CoronaVac in pediatric patients with neuromuscular diseases"

Yu Lung Lau, MBChB, MD(Hons)

Jaime S Rosa Duque, MD, PhD

### 1 Supplementary Data-Survey questions on COVID-19 vaccine hesitancy

| No. | Questions | Answer |
| --- | --- | --- |
| 1 | Patients consent | Agree / Not agree |
| 2 | Did your father receive COVID-19 vaccine? | o Yes (1)  o No (2) |
| 3 | Did your mother receive COVID-19 vaccine? | o Yes (1)  o No (2) |
| 4 | Did any other family member receive COVID-19 vaccine? | o Yes (1)  o No (2) |
| 5 | Do you need to use wheelchair? | o Yes (1)  o No (2) |
| 6 | Do you need to use Ryle’s tube or PEG tube feeding? | o Yes (1)  o No (2) |
| 7 | Do you need to use ventilator? | o Yes (1)  o No (2) |
| 8 | Have you used spinal brace / performed spinal surgery? | o Yes (1)  o No (2) |
| 9 | English full name |  |
| 10 | Age (years) |  |
| 11 | Gender | o Male (1)  o Female (2) |
| 12 | Did you get the flu vaccine last year? | o Yes (1)  o No (2) |
| 13 | Have you received the flu vaccine every year in the past three years? | o Yes (1)  o No (2) |
| 14 | Have you been diagnosed with COVID-19? | o Yes (1)  o No (2) |
| 15 | Have your family members or classmates been diagnosed with COVID-19 disease? | o Yes (1)  o No (2) |
| 16 | Have you ever been classified as a close contact of COVID-19 that needs to undergo compulsory quarantine? | o Yes (1)  o No (2) |
| 17 | Have you ever been required to compulsorily test the COVID-19 virus? | o Yes (1)  o No (2) |
| 18 | What COVID-19 vaccine have you or do you plan to receive | o BNT162b2 (1)  o Coronavac (2)  o Undecided (3) |

| 20 | Reasons to have vaccination | ▢ I think I am high risk group of COVID-19 (1)  ▢ I am worried that I will be infected (2)  ▢ I want to protect my family (3)  ▢ I want to attend religious ceremonies (4)  ▢ I want to travel (5)  ▢ I wish to return to my life before COVID-19 pandemic (6)  ▢ I am tired of the social distancing policies (7)  ▢ Because people around me received vaccination (8)  ▢ I want to resume full scale face-to-face teaching. (9)  ▢ I want to play sports at school with friends or in a competition without wearing mask (10)  ▢ I want to meet my family as I am currently living at special schools (11)  ▢ I want to play music safely (e.g. wind instruments, band, choir) (12)  ▢ I want to participate in large-scale events (e.g. open days, joint school events and Christmas balls) without wearing mask (13)  ▢ I want to study overseas (14)  ▢ I want to join study tours (15)  ▢ Socialize with large groups of friends, (e.g. dining out, karaoke, party room) (16)  ▢ Others (please specify): (17) __________________________________________________ |
| --- | --- | --- |

| 21 | Reasons for not to have vaccination? | ▢ Not allowed by my parents (1)  ▢ Medically not suitable (e.g. severe allergies, uncontrolled diabetes mellitus) (2)  ▢ I am concerned about its safety (3)  ▢ I am concerned about its efficacy (4)  ▢ I am concerned that receiving the vaccine will violate the rules of my religion (5)  ▢ I am concerned that I will have a higher chance to encounter adverse effects when compared to healthy individuals (6)  ▢ I am concerned that the vaccination would affect my current treatment (7)  ▢ COVID-19 is just a mild disease (8)  ▢ I do not know where to receive the vaccines (9)  ▢ I am difficult to access the vaccine centres (10)  ▢ I do not know whether I should receive the vaccine (11)  ▢ The community vaccine centres are inconvenient (12)  ▢ I do not know which type of COVID-19 vaccine I should receive (13)  ▢ Facemask and social distancing are sufficient (14)  ▢ Depends on the progress of the pandemic (15)  ▢ I am against vaccination (16)  ▢ Others (please specify): (17) __________________________________________________ |
| --- | --- | --- |

### 2 Supplementary Figures and Tables

#### 2.1 Supplementary Figures

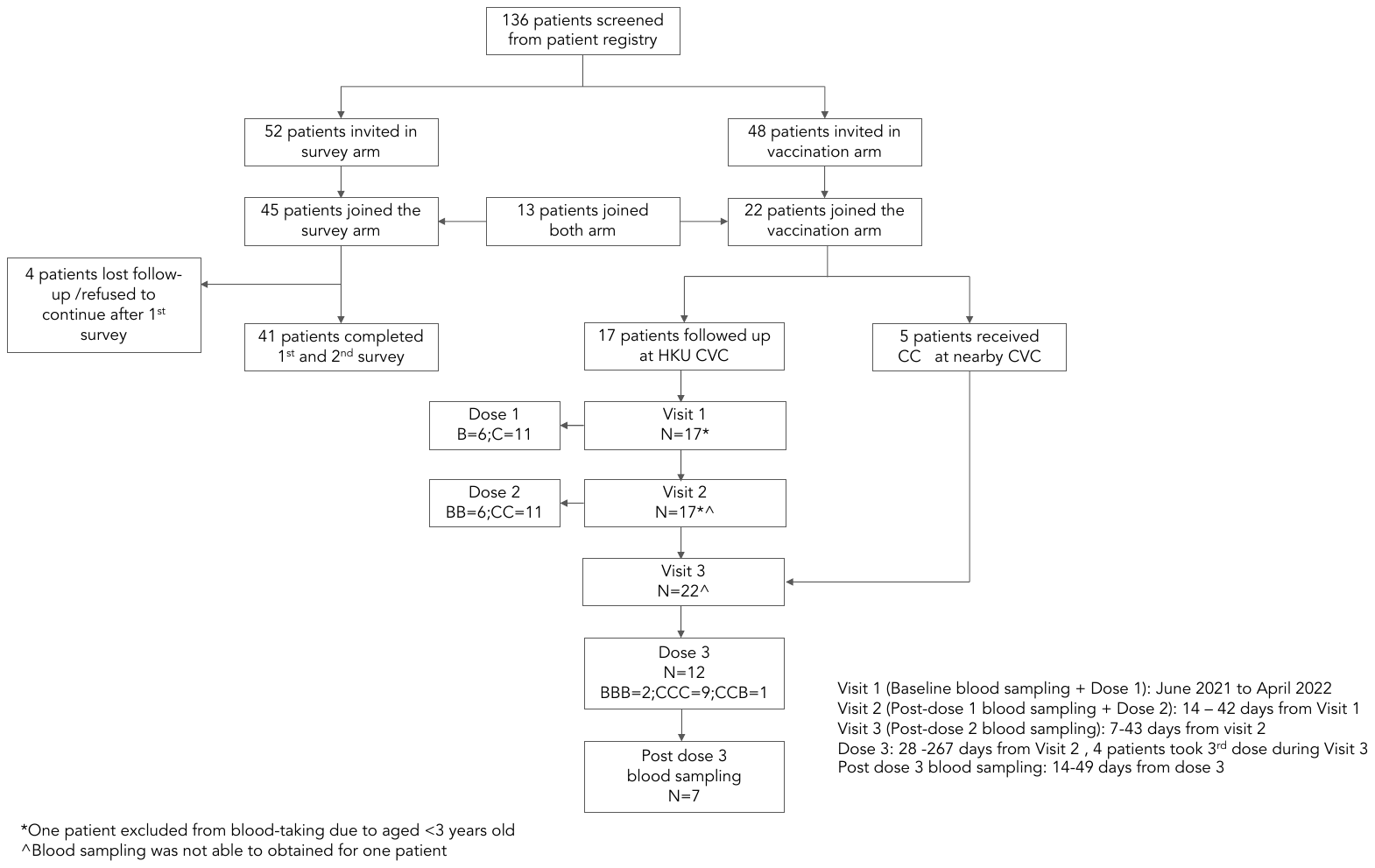

Supplementary Figure 1: Flow diagram of study participants

136 patients were screened from the patient registry. Fifty-two patients were invited to complete the hesitancy survey arm, and 48 patients were invited to join the reactogenicity and immunogenicity arm of the study. Forty-five patients completed the first hesitancy survey, and 41 (91.1%) of them completed both first and second surveys. For the reactogenicity and immunogenicity arm of the study, 22 patients joined and 17 were inoculated with BNT162b2 or CoronaVac at our HKU Community Vaccination Centers (CVCs) research site. These patients recorded adverse reactions in a 7-day diary system after vaccination for reactogenicity/safety analyses and had blood sampling. Additionally, 5 patients who received two doses of CoronaVac at nearby CVCs and had blood sampling after the second dose.

#### 2.2 Supplementary Tables

Supplementary Table 1: Demographic and clinical characteristics of the patients with neuromuscular diseases in COVID-19 vaccine hesitancy survey arm of the study

|  | N/mean | %/SD |
| --- | --- | --- |
| Demographics |  |  |
| Age (years) | 13.8 | 3.4 |
| Female | 18 | 43.9 |
| Breakdown of NMDs |  |  |
| Spinal muscular atrophy | 18 | 43.9 |
| 5q type I | 2 | 4.9 |
| 5q type II | 10 | 24.4 |
| 5q type III | 5 | 12.2 |
| Non-5q spinal muscular atrophy | 1 | 2.4 |
| Dystrophinopathy | 6 | 14.6 |
| Duchenne muscular dystrophy | 4 | 9.8 |
| Becker muscular dystrophy | 2 | 4.9 |
| Congenital myopathy | 5 | 12.2 |
| Congenital muscular dystrophy | 4 | 9.8 |
| Peripheral neuropathies | 2 | 4.9 |
| Facioscapulohumeral muscular dystrophy | 2 | 4.9 |
| Myotonic disorder | 2 | 4.9 |
| Other NMDs^ | 2 | 4.9 |
| Physical conditions |  |  |
| Wheelchair-mobile | 22 | 53.7 |
| Nasogastric tube or PEG tube feeding use | 4 | 9.8 |
| Ventilator support | 14 | 34.1 |
| Using spinal brace or had spinal surgery | 8 | 19.5 |

^Congenital myasthenic syndrome and ocular myasthenia gravis

NMDs= neuromuscular diseases, PEG= percutaneous endoscopic gastrostomy, SD= standard deviation, N= numbers

Supplementary Table 2: Intention of COVID-19 vaccination in patients with neuromuscular diseases in January and April 2022

| Intention of COVID-19 vaccination | January 2022 | April 2022 | *p*-values |
| --- | --- | --- | --- |
| Have vaccinated/ planning to vaccinate | 30 (73.2) | 40 (97.6) |  |
| Do not plan to vaccinate | 11 (26.8) | 1 (2.4) |  |
| Total | 41 | 41 | 0.003** |
| Will continue to receive future boosters, if any? |  |  |  |
| Yes | 26 (63.4) | 30 (73.2) |  |
| No | 15 (36.6) | 11 (26.8) |  |
| Total | 41 | 41 | 0.477 |

Data presented as number (%)

***p*<0.01

Supplementary Table 3: Preference on COVID-19 vaccines in patients with neuromuscular diseases

|  | | January 2022 | | | April 2022 | | |
| --- | --- | --- | --- | --- | --- | --- | --- |
|  | Types of vaccines | Planning to receive | Already vaccinated | Total | Planning to receive | Already vaccinated | Total |
|  | BNT162b2 | 5 (33.3) | 14 (93.3) | 19 (46.3) | 3 (100.0) | 17 (45.9) | 20 (48.8) |
|  | CoronaVac | 8 (53.3) | 1 (6.7) | 9 (22.0) | 0 (0.0) | 20 (54.1) | 20 (48.8) |
|  | Undecided | 2 (13.3) | 0 (0.0) | 2 (4.9) | 0 (0.0) | 0 (0.0) | 0 (0.0) |
|  | Do not plan to vaccinate | - | - | 11 (26.8) | - | - | 1 (2.4) |
|  | Total | 15 | 15 | 41 | 3 | 37 | 41 |

Data presented as number (%)

Supplementary Table 4: Demographic and clinical characteristics of the patients with neuromuscular diseases

in the COVID-19 vaccine reactogenicity and immunogenicity arm of the study

| Patient | NMD type | Age (years) | Sex | Ambulatory status | Treatment | Dosage | Dose 1 & 2 | Dose 3 | COVID-19 infection |
| --- | --- | --- | --- | --- | --- | --- | --- | --- | --- |
| 1 | CIDP | 16-20 | Male | Independent walker | IVIG | 2 g/kg/3months | BNT162b2 | N/A | N |
| 2 | DMD | 11-15 | Male | Late ambulatory | Prednisolone | 0.42 mg/kg/day | BNT162b2 | BNT162b2 | N |
| 3 | DMD | 11-15 | Male | Wheelchair mobile | Deflazacort | 0.64 mg/kg/day | BNT162b2 | N/A | N |
| 4 | CM | 11-15 | Female | Independent walker | N/A | N/A | BNT162b2 | N/A | Y, before 1^st^ dose |
| 5 | SMA | 11-15 | Male | Wheelchair mobile | Risdiplam | 0.13 mg/kg/day | BNT162b2 | N/A | N |
| 6 | SMA | 6-10 | Male | Independent walker | Nusinersen | N/A | BNT162b2 | BNT162b2 | N |
| 7 | DMD | 11-15 | Male | Late ambulatory | Prednisolone | 0.61 mg/kg/day | CoronaVac | CoronaVac | N |
| 8 | DMD | 11-15 | Male | Wheelchair mobile | Prednisolone | 0.30 mg/kg/day | CoronaVac | N/A | N |
| 9 | DMD | 0-5 | Male | Independent walker | Deflazacort | 0.64 mg/kg/day | CoronaVac | CoronaVac | N |
| 10 | SMA | 6-10 | Female | Wheelchair mobile | Nusinersen | 0.56 mg/kg/day | CoronaVac | N/A | Y, after 1^st^ dose |
| 11 | SMA | 0-5 | Male | Wheelchair mobile | Nusinersen | 0.56 mg/kg/day | CoronaVac | CoronaVac | N |
| 12^ | CM | 6-10 | Female | Wheelchair mobile | N/A | N/A | CoronaVac | BNT162b2 | N |
| 13 | DMD | 6-10 | Male | Late ambulatory | Deflazacort | 0.74 mg/kg/day | CoronaVac | CoronaVac | N |
| 14 | SMA | 0-5 | Female | Wheelchair mobile | Nusinersen | 0.83 mg/kg/4 months | CoronaVac | CoronaVac | N |
| 15 | DMD | 11-15 | Male | Wheelchair mobile | Deflazacort | 0.67 mg/kg/day | CoronaVac | N/A | Y, before 1^st^ dose |
| 16 | MD | 6-10 | Female | Independent walker | Melatonin | 5.45 mg/kg/day | CoronaVac | CoronaVac | N |
| 17 | DMD | <20 | Male | Wheelchair mobile | Prednisolone | 0.49 mg/kg/day | CoronaVac | CoronaVac | N |
| 18* | SMA | 11-15 | Male | Independent walker | Nusinersen | 0.38 mg/kg/4months | CoronaVac | N/A | N |
| 19* | DMD | 6-10 | Male | Independent walker | Prednisolone | 0.72 mg/kg/day | CoronaVac | N/A | N |
| 20* | SMA | 11-15 | Female | Wheelchair mobile | Nusinersen | 0.48 mg/kg/4months | CoronaVac | N/A | N |
| 21* | BMD | 11-15 | Male | Independent walker | N/A | N/A | CoronaVac | CoronaVac | N |
| 22* | CM | 11-15 | Female | Late ambulatory | N/A | N/A | CoronaVac | CoronaVac | N |

BMD= Becker muscular dystrophy, CIDP= chronic inflammatory demyelinating polyneuropathy, CM= congenital myopathy,

DMD= Duchenne muscular dystrophy, IVIG= intravenous immunoglobulins, MD= myotonic disorder, N/A= not applicable,

SMA= spinal muscular atrophy

^Blood samples for immunogenicity assay before the first and after the third dose of COVID-19 vaccination

*Blood samples for immunogenicity assay after the second dose of COVID-19 vaccination only

Supplementary Table 5: Comparison of immunogenicity for patients with neuromuscular diseases by corticosteroids use

| **Dosage** | **C** | **B** | **CC** | **BB** | **CCC/CCB** | **BBB** |
| --- | --- | --- | --- | --- | --- | --- |
| ELISA OD450  CS user median (N)  Non-CS user median (N)  *p*-value | 0.25 (5)  1.30 (2)  0.24 | 0.87 (2)  1.35 (3)  0.80 | 1.82 (5)  2.37 (8)  0.02* | 1.82 (2)  2.58 (3)  0.20 | 2.26 (4)  3.23 (6)  0.07 | 3.04 (1)  3.19 (1)  N/A |
| sVNT % inhibition  CS user median (N)  Non-CS user median (N)  *p* -value | 15.0 (5)  37.6 (2)  0.86 | 55.3 (2)  50.5 (3)  0.80 | 84.4 (5)  89.3 (8)  0.35 | 93.9 (2)  96.3 (3)  0.80 | 83.1 (4)  96.5 (6)  0.11 | 97.2 (1)  97.3 (1)  N/A |

Data presented as median (number of patients in group)

B, 1 dose of BNT162b2; BB, 2 doses of BNT162b2; C, 1 dose of CoronaVac; CC, 2 doses of CoronaVac; BBB, 3 dose of BNT162b2; CCC, 3 doses of CoronaVac; CCB, 2 doses of CoronaVac and 1 dose of BNT162b2.

CS= corticosteroids, ELISA= enzyme linked immunosorbent assay, N/A= Not applicable,

OD= optical density, sVNT= surrogate virus neutralization test

**p*<0.05

Supplementary Table 6: Immunogenicity of patients with neuromuscular diseases who had COVID-19 infection

| Patient No. | Vaccine | Age | Sex | COVID-19 history | Pre-dose 1  ELISA  OD450 | Pre-dose 1  sVNT % inhibition | Dose 1  ELISA  OD450 | Dose 1  sVNT % inhibition | Dose 2  ELISA  OD450 | Dose 2  sVNT % inhibition |
| --- | --- | --- | --- | --- | --- | --- | --- | --- | --- | --- |
| 4 | BB | 11-15 | F | 6 weeks before dose 1 | 0.25 | 15 | 2.56 | 86.6 | 3.14 | 97.8 |
| 10 | CC | 6-10 | F | 3 weeks after dose 1 | N/A | N/A | 1.14 | 36.1 | 2.98 | 92.4 |
| 15 | CC | 11-15 | M | 1 month before dose 1 | 0.51 | 15 | 0.53 | 15 | 1.34 | 15 |

These patients were excluded from the main immunogenicity analyses

BB, 2 doses of BNT162b2; CC, 2 doses of CoronaVac

ELISA= enzyme linked immunosorbent assay, OD= optical density, sVNT= surrogate virus neutralization test
